# The maternal anthropometry and hemoglobin status in relations to newborn birth weight among primiparous mothers at Adama Hospital Medical College, Eastern Ethiopia: a cross-sectional study

**DOI:** 10.1101/2022.05.11.22274947

**Authors:** Midekso Sento, Atoma Negera

## Abstract

**Background:** Anthropometric measurements are quantitative measurements of the human body size, shape, and nutritional status. It is a simple, inexpensive, and non-invasive approach used to identify mothers at risk of labor outcomes. On the other hand, hematological disorders are common during pregnancy with deleterious health effects on the mother and the fetus, which consequently cause fetal growth retardation and low weight.

**Objectives:** To assess the relationship between maternal anthropometry and hemoglobin status with the newborn birth weight among primiparous mothers at Adama Hospital Medical College, Eastern Ethiopia 2021.

**Methods:** An institutional-based cross-sectional study design involving 269 primiparous pregnant mothers was conducted at Adama Hospital Medical College from September 15 to October 30, 2021. The consecutive sampling technique was used till the required sample size was attained. Interviewer-administered questionnaires, instruments for anthropometric measurement, hemoglobinometer, and electronic weighing scales were used to collect the data. The data was entered into EpiData version 3.1 and exported to SPSS software version 21 for analysis. Multivariate logistic regression was used to analyze the relationship between the independent variables and the outcome variables.

**Results:** About 269 mothers and their neonates participated in this study. Multivariate logistic regression shows that maternal ANC visit less than two [AOR=4.149, 95% CI: (1.27, 13.52)], maternal height [AOR= 0.878, 95% CI: (0.806, 0.95)], and maternal Hgb <11g/dl [AOR=4.127, 95% CI: (1.63, 10.43)] were significantly associated with the occurrences of low birth weight (p<0.05).

**Conclusion:** This study has shown that maternal anthropometric characteristics and hemoglobin status were important factors associated birth weight. Health facilities should emphasize routine measurements of maternal anthropometry and hemoglobin during ANC visits in order to reduce complications related to labor outcomes.

## 1. INTRODUCTION

### Background

Anthropometric measurements are quantitative assessments of human body size and are used to assess the nutritional status of humans. Commonly used anthropometric parameters in pregnant mothers include height, weight, mid-upper arm circumference (MUAC), and foot length (1). It is a simple, inexpensive, and non-invasive approach used to identify mothers at risk of labor outcomes (2).

Researchers have used many methods, ranging from a simple measurement of maternal height to some advanced methods such as clinical or radiological pelvimetry (3). Maternal height results from the interaction of the genetic materials for growth with early-life conditions, whereas maternal weight and MUAC reflect recent nutritional status before or after conception (4). Tallness is generally considered to be advantageous, particularly concerning childbirth. Although women are genetically shorter than men, they have larger pelvic dimensions. Stature is found to be significantly related to several pelvic indices (5)..

The maternal body frame is the key determinant of neonatal anthropometry, predominantly birth weight and length, which are linked to perinatal morbidity and mortality (6). Birth weight is the major predictor of an infant’s growth and survival and is mostly affected by the mother’s health and nutritional status before conception and throughout pregnancy. It is classified as normal birth weight, low birth weight, and macrosomia (7).

The World Health Organization (WHO) defines low birth weight as a weight at birth of less than 2,500 grams, regardless of gestational age (8). It leads to stunted growth, a higher risk of mortality and morbidity, affects organ development, and a higher risk of chronic disorders in later life (7). This represents the main cause of child mortality and morbidity. Its consequences are not only limited to early life stages but also happen during adulthood. There is increasing evidence that children born with LBW have poor developmental outcomes (9).

During pregnancy, an increase in plasma volume exceeds the increase in red cell volume, which causes a physiological hemodilution, resulting in a reduced hemoglobin concentration (10). Maternal anemia is one of the common hematological disorders that may occur during pregnancy and is associated with fetal and neonatal morbidity and mortality, which also accounts for 20% of maternal deaths in developing countries (11).

Anthropometry and the hemoglobin status of mothers are important factors that influence birth weight; in turn, weight at birth is related to neonatal outcomes (12). Low birth weight (LBW) is the leading cause of neonatal morbidity and mortality in Africa (13). Infants weighing less than 2,500 grams are 20 times more likely to die than bigger babies, and low birth weights contribute to poor health outcomes (14). Low birth weight continues to be a major public health problem globally and is associated with a variety of both short and long-term outcomes. Low birth weight (LBW) is estimated to account for 15% to 20% of all births worldwide, which accounts for more than 20 million births a year. Based on the report of WHO, the regional estimates of LBW include 9% in Latin America, 28% in South Asia, and 13% in sub-Saharan Africa (15). And in Ethiopia, the national pooled prevalence of low birth weight (LBW) was 17.3% (16)

Globally, 528.7 million (29.4%) women of reproductive age have hemoglobin (Hb) concentrations of less than 11 g/dl. Rates of low hemoglobin are highest in low-resource countries, especially in central and western Africa, where 48% of reproductive-age women and 56% of all pregnant women are reported to have low hemoglobin (18). Maternal low hemoglobin constitutes a major public health problem in developing countries. Maternal anemia in pregnancy is common and has several deleterious effects on the health of the mother and the fetus, and is an important risk factor for low birth weight (LBW) (19). Several studies conducted in various parts of the world found that maternal anthropometry and hemoglobin status are important factors influencing labor outcomes. Understanding the relationship between maternal anthropometry and hemoglobin status with newborn birth weight aids in enforcing early management and follow-up in order to prevent both short and long-term complications. Besides, this study provides information on the associations between maternal anthropometry and hemoglobin status with the newborn birth weight among primiparous mothers at Adama Hospital Medical College.

## 2. METHODS AND MATERIALS

### 2.1. Study area and period

The study was conducted at Adama Hospital Medical College (AHMC) from September 15 to October 30, 2021. The hospital is one of the tertiary and teaching hospitals in the Oromia region, 99 km east of Addis Ababa. AHMC was constructed by American Mennonite missionaries in 1946. It is serving over five million catchment populations and is a referral site for neighboring zone and regions such as part of Amhara and Afar regions. There are 11 obstetricians, 39 obstetrics and gynaecology (OB/GYN) residents, and 46 midwives who are currently providing maternal and child health (MCH) related services. Based on the report of the AHMC record liaison office, there were about 9054 pregnant mothers who gave birth at AHMC in 2013 E.C. Among these, 4022 were primiparous, and on average, about 335 primiparous gave birth each month at AHMC.

### 2.2. Study design

An institutional-based cross-sectional study design was conducted at AHMC.

### 2.3. Population

#### 2.3.1 Source population

All primiparous mothers presented for delivery at Adama Hospital Medical College.

#### 2.3.2 Study population

Primiparous mothers who fulfilled the inclusion criteria at Adama Hospital Medical College.

### 2.4. Inclusion and Exclusion Criteria

#### 2.4.1 Inclusion Criteria

➤ Full-term pregnancy
➤ Primiparous who delivered singleton live birth

#### 2.4.2 Exclusion Criteria

➤ Primiparous with multiple pregnancies, medical disorders of pregnancy, such as gestational hypertension, diabetes, oligohydramnios, and mothers suspected of COVID19.
➤ Newborn with congenital anomalies, and stillbirth.

### 2.5. Sample size determination

The sample was determined using the formula for single population proportions, taking proportions of 0.21 of low birth weight under the assumption of a 95% confidence interval (Z=1.96) and a margin of error of 0.05 (31). Hence, the number of deliveries at the hospital in 2013 EC was 9054, which is less than 10,000. The sample size correction formula was applied, and finally, after accounting for a 10% non-response rate, the sample size was 273.

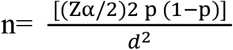

Where:

n= minimum sample size required for the study

Z (α/2)2 = standard normal distribution (Z=1.96), CI of 95% = 0.05

P= proportion of low birth weight 0.21 (31)

d=Absolute precision or tolerable margin of error= 5 %(0.05)

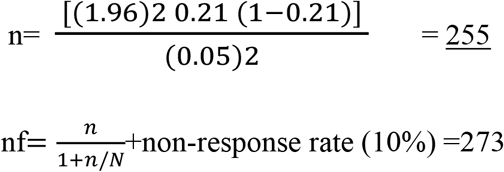

### 2.6. Sampling technique

A consecutive sampling method was used to collect the data during the study period. All primiparous mothers presented for delivery who met the inclusion criteria were consecutively recruited until the required sample size was attained.

### 2.7. Operational definitions and definition of terms

#### Primiparous

Women who have never given birth to a child other than the current one

#### Term pregnancy

Pregnancy with GA between 37 -42 weeks (23).

#### Birth weight

Classified as birth weight less than 2.5kg and greater or equal to 2.5kg

#### Maternal anemia

Maternal Hgb concentration less than 11g/dl (32).

#### Low birth weight

Newborn with birth weight less than 2.5kg

#### Height

Measured in the standing position following standards of measuring height by using a stadiometer (mothers stood next to a wall with their feet and knees together, knees straight, heels, legs, hip, shoulders, back of the head parallel to the wall, their body completely flat and stretched, hands hanging on both sides, and looking straight ahead and standing height was measured in centimeter (cm) (17).

#### Weight

Measured by using a weight scale with a light cloth and without shoes after delivery in kilogram.

#### MUAC

Measured in the arm at the level, midway between acromion and olecranon processes in centimeter (cm).

#### Foot length

Is the distance from heel to the tip of the longest toe measured in cm (23).

### 2.8. Data Collection Methods

#### 2.8.1. Data Collection Instruments

An interviewer-based structured questionnaire prepared in English and translated into the local language, Afan Oromo was used to obtain maternal socio-demographic information. The questionnaire comprised three parts: socio-demographic factors, maternal characteristics, and newborn data. The questionnaire comprised a total of fourteen questions. The first part was regarding the socio-demographic characteristics of the participants, which contained five questions; the second part was about maternal anthropometry, hemoglobin status, and mode of delivery, which contained seven questions; and the third part was newborn data, which contained two questions.

A weight scale, MUAC tape, stadiometer, and wooden centimeter were used to obtain maternal anthropometry. A portable hemoglobinometer was used to measure maternal hemoglobin status. And an electronic weighing scale was used to measure the newborn weight.

#### 2.8.2. Data Collectors and Data Collection Procedures

The data collection was conducted by four experienced BSc holders’ midwives who could speak and understand the local language. The principal investigators assisted and coordinated the overall data collection process. To obtain maternal socio-demographic information, an interviewer-administered structured questionnaire was used. Maternal height was measured in the standing position following the standard of measuring height. It was measured after delivery with a stadiometer that had a movable headpiece perpendicular to a well-calibrated meter scale and was recorded to the nearest cm. The mothers were weighed in light clothes and without shoes. The weighing scale was checked for zero adjustments before each use, and the weighting scale was placed on a flat and hard surface.

The mid-upper arm circumference (MUAC) was measured in the arm at the level midway between the acromion and olecranon processes to the nearest cm. Foot length is measured by a wooden centimeter from the heel to the tip of the longest toe. All maternal anthropometric measurements were taken three times, and the average was taken (17). Maternal hemoglobin (Hgb) was determined using a portable hemoglobinometer, the HemoCue Hgb analyzer (HemoCue Hb 201+, Sweden), by taking a drop of blood and puncturing the tip of the middle finger of the mother with a sterile lancet. The procedure of maternal hemoglobin was done following the standard guidelines at the labor ward before the delivery of the newborn (33). Newborn weight was measured immediately after birth by using electronic weighing scales in grams. All maternal anthropometrics were measured within 24 hrs. of delivery.

### 2.9. Data processing and analysis

The data was checked for completeness manually and entered into the EpiData version 3.1 software package. For further analysis, the data was exported to the Statistical Package for Social Science (SPSS) version 21 software. A descriptive analysis was performed for each of the independent variables, and the results were presented in the form of tables and graphs. The data was fitted with binary logistic regression. Bivariable analysis was done to test for the relationship between dependent and independent variables and model fitness was checked by using the Hosmer-Lemeshow goodness of fit test. The variance inflation factor (VIF) was used to test for multicollinearity. The logistic regression results were presented as adjusted odds ratio (AOR). Finally, those variables with a P-value of less than 0.05 with a 95% confidence interval in the multivariable analysis were considered statistically significant predictor variables of the birth weight. All variables were entered into a multivariable binary logistic regression analysis to identify factors independently associated with birth weight.

### 2.10. Data quality management

To assure the quality of the data, one-day training was given for data collectors on the objective of the study, inclusion and exclusion criteria, contents of the questionnaires, procedures for anthropometric measurements, and ethics during data collection. The collection of data was checked by the investigator daily for any incompleteness and/or inconsistency. A pre-test was done to assess the understanding level of the data collectors and the integrity of the questionnaire among 5% (14) of the sample size at Adama General Hospital Medical College. Each questionnaire was identified by the ID code given to it.

### 2.11. Ethical consideration

Ethical clearance to conduct the study was obtained from the Jimma University Institute of Health’s Institutional Review Board with a reference number of IHRPGS/521/2021. A written letter of permission for data collection was provided to Adama Hospital Medical College administrative office, department of obstetrics and gynecology. All data collectors who participated in the study were recruited by staff working in the obstetric ward while simultaneously conducting their routine duty in the hospital.

The study title, purpose, procedure, duration, possible risks, and benefits of the study were clearly explained to the study participants. Then, individual written informed consent was taken from the respondents, and they were assured of confidentiality by excluding their names during the period of data collection. They were informed that they have the full right to refuse to participate and/or withdraw from the study at any time if they have any difficulty. Regarding newborns in which LBW was diagnosed; communication with the midwives in charge was made for further assessment and management. Prevention of COVID-19 was considered by wearing personal protective equipment and applying necessary infection prevention techniques during all steps of data collection.

## 3. RESULTS

### 3.1 Maternal socio-demographic characteristics

A total of 269 primiparous mothers and their newborn neonates participated in this study, which yields a response rate of 98.5%. Table 2 presents the socio-demographic characteristics of primiparous mothers who gave birth at Adama Hospital Medical College. The mean of maternal age was 23.95±3.42. More than three forth 210 (78.1%) of the participants were urban in their place of residence. Besides, about four in ten mothers 115 (42.7%) had a secondary level of education. Nearly about eight in ten 224 (83.3%) mothers were unemployed. According to this study, more than half 144 (53.5%) of the participants had at least four ANC visits. (Table 1)

**Table 1:** Socio-demographic characteristics of primiparous mothers who gave birth at Adama Hospital Medical College 2021(n=269)

| Variables | Categories | Mode of delivery |  | Total |
| --- | --- | --- | --- | --- |
|  |  | VD Mean/N (%) | CS Mean/N (%) | Mean/ N (%) |
| Age | --- | 24.08±3.28 | 23.59±3.80 | 23.95±3.42 |
| Residence | Rural | 45(16.7%) | 14(5.2%) | 59(21.9%) |
|  | Urban | 154(57.2%) | 56(20.8%) | 210(78.1%) |
| Educational level | No education | 14(5.2%) | 7(2.6%) | 21(7.8%) |
|  | Primary(1-8) | 57(21.2%) | 19(7%) | 76(28.2%) |
|  | Secondary(9-12) | 83(30.8%) | 32(11.9%) | 115(42.7%) |
|  | Higher level(>12) | 45(16.7%) | 12(4.5%) | 57(21.2%) |
| Occupational status | Unemployed | 163(60.6%) | 61(22.7%) | 224(83.3%) |
|  | Employed | 36(13.4%) | 9(3.3%) | 45(16.7%) |
| ANC visit | <2 visit | 12(4.46%) | 10(3.7%) | 22(8.1%) |
|  | 2-3 visit | 78(28.9%) | 25(9.3%) | 103(38.3%) |
| | ≥4 visit | 109(40.5%) | 35(13.0%) | 144(53.5%) |

VD= Vaginal delivery, CS= Cesarean section, N= Number/frequency

**Table 2:** Mean of maternal characteristics in association to birth weight of neonates at Adama Hospital Medical College 2021(n=269)

| Variables | BW <2500g | BW ≥2500g | Total mean± SD |
| --- | --- | --- | --- |
|  | mean±SD | mean±SD |  |
| Maternal Age | 22.48±2.70 | 24.32±3.49 | 23.95±3.42 |
| Height | 159.12±3.01 | 161.54±5.12 | 161.16±4.92 |
| Weight | 51.72±3.82 | 60.53±8.98 | 59.12±8.97 |
| Foot length | 22.57±1.412 | 23.062±1.044 | 22.96±1.141 |
| MUAC | 22.67±1.38 | 25.03±2.51 | 24.66±2.52 |
| Hemoglobin | 11.48±1.03 | 12.64±1.41 | 12.46±1.43 |

**BW**= birth weight, **SD**= standard deviation

The distribution of newborn sex among neonates delivered at Adama Hospital Medical College. Out of the total neonates who participated in this study, 159 (59.1%) were male and 110 (40.9%) were female. (Figure 1). And Out of the total newborns who participated in this study, 215 (80%) had birth weights greater or equal to 2500g and 54 (20%) had birth weights less than 2500g. (Figure 2)

**Figure 1:**
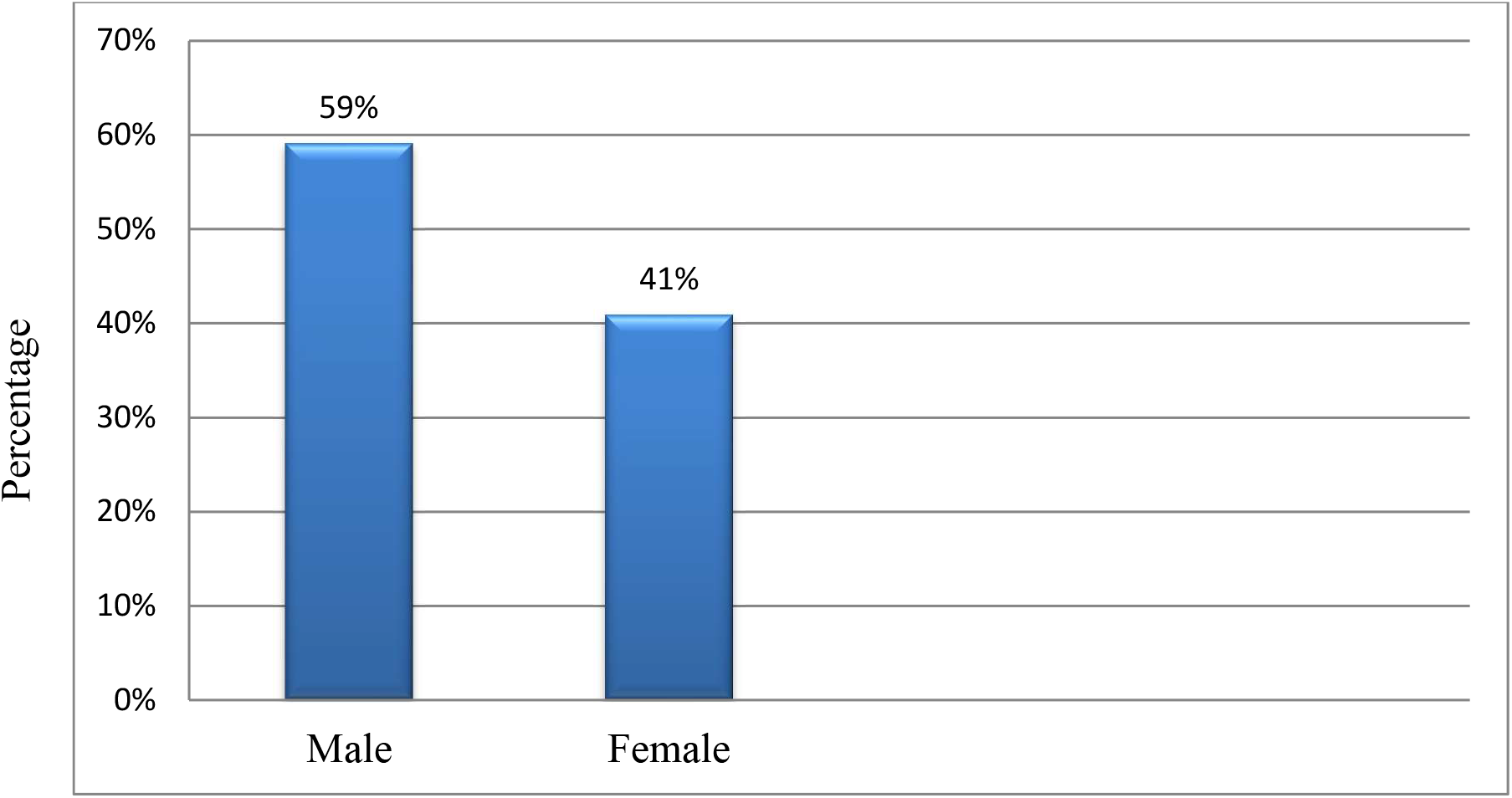
Sex of newborn among neonates delivered by primiparous mothers at Adama Hospital Medical College 2021 (n=269)

**Figure 2:**
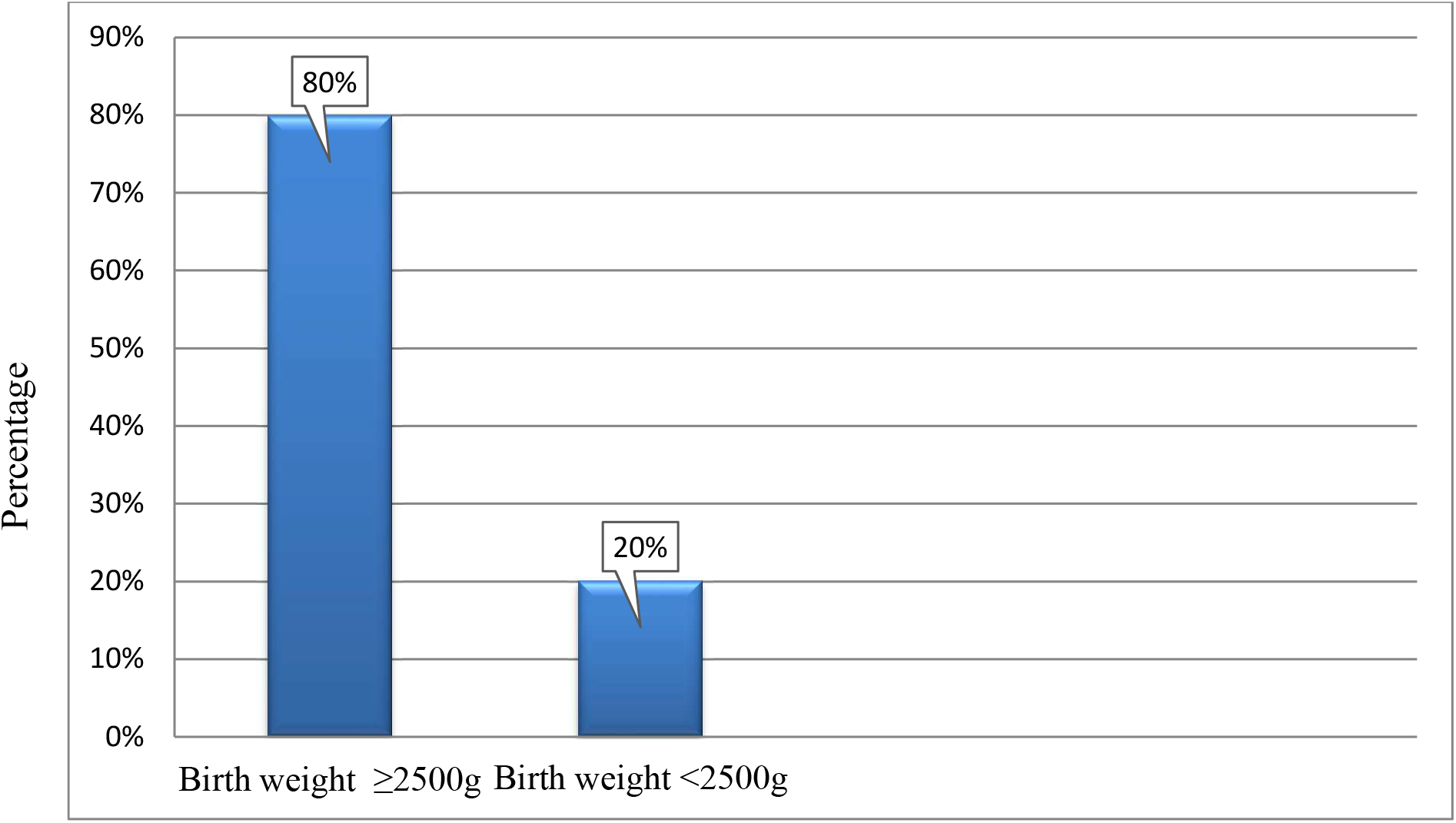
Birth weight status among newborns delivered by primiparous mothers at Adama Hospital Medical College 2021 (n=269)

### 3.2 Mean of maternal characteristics in association to birth weight of newborns

Table 2 shows the mean comparison of maternal characteristics in association with the birth weight of neonates at Adama Hospital Medical College. In comparison to the mothers who delivered neonates with a birth weight of less than 2500g, mothers who delivered neonates with a birth weight greater than or equal to 2500g had a higher mean of maternal age, maternal foot length, maternal height, maternal weight, maternal MUAC, and maternal hemoglobin. (Table 2)

### 3.3 Independent predictors associated with low birth weight (LBW)

To identify the association between low birth weight (LBW) and predictive variables, bivariable binary logistic regression analysis was first done for all independent variables. Table 6 presents independent predictors associated with low birth weight (LBW) at Adama Hospital Medical College. All important variables were entered into the multivariable binary logistic regression by ‘ENTER METHOD’. Model fitness was checked by the Hosmer-Lemeshow goodness of fit test. Accordingly, the result was 0.713 for maternal characteristics and newborn sex in association to birth weight, and then the multivariable binary logistic regression analysis showed maternal height, maternal hemoglobin status, and maternal ANC visit of less than two were associated with low birth weight (LBW).

When compared to mothers who had four or more ANC visits, primiparous mothers with fewer than two ANC visits increased the occurrence of low birth weight (LBW) by 4.149 [AOR = 4.149, 95% CI: (1.27, 13.52)] folds. As maternal height increased by one unit, the likelihood of low birth weight (LBW) decreased by 12.2% [AOR = 0.878, 95% CI: (0.806, 0.95)]. Besides, the odds of having a low birth weight (LBW) were 4.127 [AOR = 4.127, 95% CI; (1.63, 10.43)] times higher among primiparous mothers with Hgb<11g/dl compared to mothers with Hgb ≥11g/dl. (Table 3).

**Table 3:** Independent predictors associated with low birth weight (LBW) at Adama Hospital Medical College 2021(n=269)

| Variables | BW<2500g<br>Mean±SD/N<br>(%) | COR(95%CI) | AOR(95%CI) |
| --- | --- | --- | --- |
| Maternal Age | 22.48±2.70 | 0.829(0.746, 0.92) | 0.902(0.792, 1.027) |
| Maternal Height | 159.12±3.01 | 0.879(0.822, 0.94) | 0.878(0.806, 0.95)* |
| Maternal Weight | 51.72±3.82 | 0.971(0.875, 0.96) | 0.972(0.899, 1.05) |
| Maternal Foot length | 22.57±1.412 | 0.684(0.522, 0.89) | 0.900(0.648, 1.252) |
| Maternal MUAC | 22.67±1.38 | 0.743(0.640, 0.86) | 0.910(0.697, 1.189) |
| Maternal Hgb status |  |  |  |
| <11g/dl | 14(5.2%) | 4.076(1.860, 8.93) | 4.127(1.63, 10.43)* |
| ≥11g/dl | 40(14.8%) | 1 | 1 |
| Residence |  |  |  |
| Rural | 37(13.7%) | 1.893(0.972, 3.68) | 2.19(0.965, 4.97) |
| Urban | 17(6.3%) | 1 | 1 |
| Educational level |  |  |  |
| No | 6(2.2%) | 2.133(0.653, 6.97) | 5.520(0.830, 36.73) |
| education | 20(7.4%) | 1.905(0.793, 4.57) | 2.693(0.574, 12.64) |
| Primary | 19(7%) | 1.056(0.444, 2.50) | 1.777(0.410, 7.695) |
| Secondary | 9(3.3%) | 1 | 1 |
| Higher level |  |  |  |
| Occupational status |  |  |  |
| Unemployed | 46(17.1%) | 1.195(0.521, 2.74) | 0.233(0.048, 1.123) |
| Employed | 8(2.9%) | 1 | 1 |
| ANC visit |  |  |  |
| <2 visits | 10(3.71%) | 4.621(1.78, 11.99) | 4.149(1.27, 13.52)* |
| 2-3 visits | 22(8.17%) | 1.506(0.783, 2.83) | 0.833(0.386, 1.79) |
| ≥4 visit | 22(8.17%) | 1 | 1 |
| Sex of new born |  |  |  |
| Male | 39(14.5%) | 2.058(1.071, 3.95) | 2.12(0.975, 4.60) |
| Female | 15(5.6%) | 1 | 1 |

\*P-value <0.05, AOR= Adjusted odds ratio, COR= Crude odds ratio CI= Confidence interval, 1= Reference

## 4. DISCUSSION

This study assessed the relationship between maternal anthropometry and hemoglobin status with the newborn birth weight among primiparous mothers at Adama Hospital Medical College. Accordingly, maternal ANC visits less than two, maternal height and maternal hemoglobin status of the mothers were significantly associated with low birth weight (LBW).

Our study found that low birth weight (LBW) was higher among mothers with an ANC visit of less than two when compared to four or more ANC visits. This finding is in line with studies conducted in Thailand (21),(22). The possible reason for the occurrence of low birth weight among mothers with low antenatal care (ANC) visits might be due to the importance of ANC to the health of pregnant mothers. Antenatal visits of pregnant mothers are very important as they provide chances for monitoring fetal wellbeing and allow timely intervention for feto-maternal protection. Little ANC could increase prenatal feto-maternal complications. And perhaps due to being well aware of mothers with high ANC visits regarding diet during pregnancy, as she was well counselled by her health care provider during visits.

This study also found that there was a negative relationship between maternal height and the likelihood of low birth weight. This finding is in agreement with studies conducted in Sudan (13), Japan (24), UK (3), India (25), and Iran (20). Mothers who are short in height may have a narrow pelvis, resulting in limited intrauterine space. This may restrict intrauterine fetal growth. However, this is insufficient to explain the mechanism of the relationship between maternal height and low birth weight (LBW). There may be other reasons why the growth of newborns is restricted (34). However, the present finding is inconsistent with the study done in India (6), which stated that there is no relationship between maternal height and birth weight. The possible reasons might be the large sample size (n = 520) and the difference in study design (prospective) in the study conducted in India.

Additionally, our study found that there was an indirect relationship between maternal mid-upper arm circumference (MUAC) and the odds of low birth weight, even though it was not statistically significant. This result is in line with the studies conducted in Ethiopia (9), Nigeria (26), and India (6). The possible reason for their indirect association may be due to the effects of current maternal nutrition on the birth weight of the unborn fetus. As maternal MUAC is the key indicator of current maternal nutritional status, mothers with high MUAC measurements are well-nourished and have a high chance of having a baby with a higher birth weight when compared to mothers with low MUAC measurements. This might be due to the high flow of reserved nutrition from mothers with high MUAC measurements through the placenta to the unborn fetus.

Our study further revealed that low maternal hemoglobin levels were associated with low birth weight. The finding in our study is in accordance with studies conducted in Addis Ababa (9), India (29), Brazil (27), and Turkey (28), which show the association between maternal hemoglobin and birth weight. The reason for their association might be the transfer of a sufficient amount of oxygen from mothers with a higher level of hemoglobin to the unborn fetus through the placenta than from mothers with lower hemoglobin levels. Poor placental development due to low hemoglobin levels in early pregnancy, resulting in insufficient oxygen supply, has been proposed as a mechanism causing low birth weight (30)

## Conclusion

The current study attempted to identify the relationship between maternal anthropometry and hemoglobin status with newborn birth weight. Measurements of maternal anthropometry and hemoglobin represent a simple method to detect primiparous mothers at risk for labor outcomes. Low birth weight (LBW) is more likely to occur among primiparous mothers with fewer than two ANC visits. As maternal height increased, the likelihood of low birth weight (LBW) decreased. Besides, the odds of having a low birth weight (LBW) were higher among mothers with lower hemoglobin.

### Recommendations

Based on the study findings, the following recommendations were forwarded for the policy makers, national, regional and zonal health bureaus, for health care workers, and researchers. Maternal anthropometry is a very useful tool in identifying mothers at risk of labor outcomes. Therefore, routine measurements of maternal anthropometry and hemoglobin during an ANC visit. Community-level health education should be provided for women of childbearing age regarding the association between maternal anthropometry and hemoglobin level with labor outcomes. Longitudinal studies in multicenter with larger sample sizes that incorporate pelvic diameters are needed. A community-based study is recommended during which anthropometric parameters like weight can be taken before pregnancy and followed up throughout the pregnancy.

### Limitation of the study

Anthropometric measurements such as maternal weight are more reliable before conception. However, in our environment, women commonly present to health facilities only when they are advanced in pregnancy and there may be no record of their pre-pregnancy weight. This study was conducted in a single center and used a relatively small sample size. Another limitation of this study is we didn’t measure Pelvic diameter (pelvimetry).

## Data Availability

data available on reasonable request from the corresponding author

## Acknowledgment

We would like to extend our deepest heartfelt gratitude to all study participants for their voluntary participation, patience and for providing essential information. We also grateful to data collectors and supervisors for their responsible data collection during the data collection period. Lastly, we like to thank Adama Hospital Medical College administration and health professionals for their cooperation and support during data collection.

## Contributorship

Conception and design of the study: MS and AN. Conduct of the study: MS. Analysis and interpretation of data: MS and AN. Drafting the manuscript and revising it critically: MS and AN. All authors have given final approval for the manuscript to be published.

## Availability of data and materials

Datasets used and/or analyzed during the current study are available from the corresponding author on reasonable request.

## Acronyms and Abbreviations

AHMC: Adama Hospital Medical College
ANC: Antenatal care
AOR: Adjusted odds ratio
BW: Birth weight
CI: Confidence Interval
COR: Crude Odds ratio
CPD: Cephalo-pelvic disproportion
CS: Cesarean section
HBG: Hemoglobin
IRB: Institutional Review Board
LBW: Low Birth Weight
MUAC: Mid-upper arm circumference
VD: Vaginal delivery
WHO: World Health Organization

## REFERENCES

1. Cashin K, Oot L. Guide to Anthropometry: A Practical Tool for Program Planners, Managers, and Implementers. Food Nutr Tech Assist III Proj (FANTA)/ FHI 360. 2018;1–231.

2. Elshibly EM, Schmalisch G. Relationship between maternal and newborn anthropometric measurements in Sudan. Pediatr Int. 2009;51(3):326–31.

3. Prasad M, Al-Taher H. Maternal height and labour outcome. J Obstet Gynaecol (Lahore). 2002;22(5):513–5.

4. Horta BL, Barros FC, Lima NP, Assunção MCF, Santos IS, Domingues MR, et al. Maternal anthropometry: Trends and inequalities in four population-based birth cohorts in Pelotas, Brazil, 1982-2015. Int J Epidemiol. 2019;48:I26–36.

5. van Roosmalen J, Brand R. Maternal height and the outcome of labor in rural Tanzania. Int J Gynecol Obstet. 1992;37(3):169–77.

6. Patra S SG. association between maternal anthropometry and birth outcome. index copernicus Int. 2017;6.2.

7. Devaki G, Shobha R. Maternal anthropometry and low birth weight: A review. Biomed Pharmacol J. 2018;11(2):815–20.

8. Alemu A, Abageda M, Assefa B, Melaku G. Low birth weight: Prevalence and associated factors among newborns at hospitals in kambata-tembaro zone, southern Ethiopia 2018. Pan Afr Med J. 2019;34:1–8.

9. Alemu B, Gashu D. Early Human Development Association of maternal anthropometry, hemoglobin and serum zinc concentration during pregnancy with birth weight. Early Hum Dev. 2020;142(October 2019):104949.

10. Alizadeh L, Raoofi A, Salehi L, Ramzi M. Impact of maternal hemoglobin concentration on fetal outcomes in adolescent pregnant women. Iran Red Crescent Med J. 2014;16(8):1–5.

11. Nair M, Gireesh S, Yakoob R, Cherian NC. Effect of maternal anaemia on birth weight of term babies. 2018;5(3):1019–22.

12. Thame M, Osmond C, Bennett F, Wilks R, Forrester T. Fetal growth is directly related to maternal anthropometry and placental volume. 2004;894–900.

13. Elshibly EM, Schmalisch G. The effect of maternal anthropometric characteristics and social factors on gestational age and birth weight in Sudanese newborn infants. BMC Public Health. 2008;8:1–7.

14. United Nations Children’s Fund and World Health Organization, Low Birthweight: Country, regional and global estimates. UNICEF NY. low birth weight global estimate. 2004.

15. Ge H, Liu W, Li H, Zhang M, Zhang M, Liu C, et al. The association of vitamin D and vitamin E levels at birth with bronchopulmonary dysplasia in preterm infants. Pediatr Pulmonol. 2021;

16. Endalamaw A, Engeda EH, Ekubagewargies DT, Belay GM, Tefera MA. Low birth weight and its associated factors in Ethiopia: A systematic review and meta-analysis. Ital J Pediatr. 2018;44(1):1–12.

17. Solomon D, Dirar A, Getachew F. Age, Anthropometric Measurements and Mode of Delivery among Primigravidae Women at Addis Ababa Governmental Hospitals, Ethiopia. J Women’s Heal Care. 2018;07(01):1–6.

18. Ali SA, Tikmani SS, Saleem S, Patel AB, Hibberd PL, Goudar SS, et al. Hemoglobin concentrations and adverse birth outcomes in South Asian pregnant women: findings from a prospective Maternal and Neonatal Health Registry. Reprod Health. 2020;17(Suppl 2):1–13.

19. Gnanasekaran SV.JR.RR. Study on the effect of maternal anemia on birth weight of term neonates among rural population India. Int J Contemp Pediatr. 2019;6(3):1255.

20. Tabrizi FM, Saraswathi G. Maternal anthropometric measurements and other factors : relation with birth weight of neonates. 2012;6(2):132–7.

21. Siramaneerat I, Agushybana F, Meebunmak Y. Maternal Risk Factors Associated with Low Birth Weight in Indonesia. Open Public Health J. 2018;11(1):376–83.

22. Oulay L, Laohasiriwong W, Phajan T, Assana S, Suwannaphant K. Effect of antenatal care on low birth weight prevention in Lao PDR: A case control study [version 1; peer review: 1 approved with reservations, 1 not approved]. F1000Research. 2018;7.

23. Solomon D. Validation of Maternal Anatomical Anthropometric Measurements to Predict Cephalopelvic Disproportion Among primigravid Women Visiting Governmental Hospitals in Addis Ababa, Ethiopia. 2017;

24. Kuritani Y, Hayashi S, Yamamoto R, Mitsuda N, Ishii K. Association between maternal height and mode of delivery in nulliparous Japanese women. J Obstet Gynaecol Res. 2020;46(12):2645–50.

25. Krishna C. Maternal Anthropometry and it’s Relationship to Birth Weight. Int J Prev Curative Community Med. 2018;04(04):48–53.

26. CU Onubogu I, Egbuonu EU. Size of Term Singleton South-East Nigerian Newborn Infants. 2017;852–9.

27. Claudia A, Godoy M, Id F, Gomes-filho IS, Orrico S, Carvalho E, et al. Maternal anemia and birth weight : A prospective cohort study. 2019;1–14.

28. Dane B, Arslan N, Batmaz G, Dane C. Does maternal anemia affect the newborn ? 2013;(June 2009):195–9.

29. Tabrizi FM, Saraswathi G. Maternal anthropometric measurements and other factors: Relation with birth weight of neonates. Nutr Res Pract. 2012;6(2):132–7.

30. Levy A, Fraser D, Katz M, Mazor M, Sheiner E. Maternal anemia during pregnancy is an independent risk factor for low birthweight and preterm delivery. 2005;122.

31. Sema A, Tesfaye F, Belay Y, Amsalu B, Bekele D, Desalew A. Associated Factors with Low Birth Weight in Dire Dawa City, Eastern Ethiopia: A Cross-Sectional Study. Biomed Res Int. 2019;2019.

32. Ifa D. Prevalence of anemia and associated factors among term neonate. 2021;

33. Hb H, Record R. HemoCueHb201+: Patient Test Procedure Purpose: Required Materials: Procedure 1-4. 2017;1–4.

34. Inoue S, Naruse H, Yorifuji T, Kato T, Murakoshi T, Doi H, et al. Association between short maternal height and low birth weight: A hospital-based study in Japan. J Korean Med Sci. 2016;31(3):353–9.

